# COVID-19 TARRACO Cohort Study: Development of a predictive prognostic rule for early assessment of COVID-19 patients in primary care settings

**DOI:** 10.1101/2020.12.11.20247932

**Authors:** Angel Vila-Corcoles, Eva Satue-Gracia, Angel Vila-Rovira, Cinta de Diego-Cabanes, Maria Jose Forcadell-Peris, Imma Hospital-Guardiola, Olga Ochoa-Gondar

## Abstract

**PURPOSE:** Clinical course in COVID-19 patients is uncertain. This study investigated possible early prognostic factors among middle-aged and older adult and explored prognostic rules stratifying risk of patients.

**METHODS:** Community-based retrospective cohort study that included 282 community-dwelling symptomatic patients ≥50 years with laboratory-confirmed COVID-19 (hospitalised and/or outpatient) during March-June 2020 in Tarragona (Southern Catalonia, Spain). Relationship between demographics, pre-existing comorbidities and early symptomatology (first 5-days) and risk of suffering critical outcome (ICU-admission/death) across clinical course was evaluated by logistic regression analyses, and simple predictive models were developed.

**RESULTS:** Of the 282 cases (mean age: 65.9 years; 140 men), 154 (54.6%) were hospitalised (30 ICU-admitted) and 45 (16%) deceased. In crude analyses, increasing age, male sex, some comorbidities (renal, respiratory or cardiac disease, diabetes and hypertension) and symptoms (confusion, dyspnea) were associated with an increased risk to suffer critical outcome, whereas other symptoms (rinorrhea, myalgias, headache, anosmia/disgeusia) were related with reduced risk. After multivariable-adjustment only age/years (OR: 1.04; 95% CI: 1.01-1.07; p=0.004), confusion (OR: 5.33; 95% CI: 1.54-18.48; p=0.008), dyspnea (OR: 5.41; 95% CI: 2.74-10.69; p<0.001) and myalgias (OR: 0.30; 95% CI: 0.10-0.93; p=0.038) remained significantly associated with increased or reduced risk. A proposed CD65-M prognostic rule (including the above mentioned 4 variables) showed a good correlation with the risk of suffering critical outcome (area under ROC curve: 0.828; 95% CI: 0.774-0.882).

**CONCLUSION:** Clinical course of COVID-19 is early unpredictable, but simple clinical tools as the proposed CD65-M rule (pending external validation) may be helpful assessing these patients in primary care settings.

## INTRODUCTION

The novel coronavirus SARS-COV-2 infection is causing the greatest infectious pandemic disease (COVID-19) in this century, with more than 60 million infected persons to date.(1-3) Despite some time elapsed since pandemic started, population-based data about susceptibility and risk factors for COVID-19 is limited. There are numerous studies reporting clinical features and outcomes among hospitalised patients with severe COVID-19, but community- or population-based data including the overall spectrum of disease (i.e., patients with mild or moderate symptoms managed as outpatient or in ambulatory basis) is scarce.(1,3)

It seems well established that the most severe and fatal cases, although they can occur in young and “healthy” people, are more frequent in older people and / or with comorbidities, like cardiovascular disease, hypertension, diabetes, chronic respiratory disease or chronic kidney disease.(4-6) Smoking, obesity, male sex, socioeconomic deprivation and ethnicity were also associated with greater severity/lethality.(7,8) Lastly, multiple laboratory findings, viral and genetics factors may be associated with worse outcomes.(9,10)

Several prognostic rules/algorithms have been reported by combining demographic, clinical, radiological and laboratory data.(11-16) However, since a family physician point of view, the majority of predictive markers are not very useful in primary care settings because most of them requires radiological and/or analytical data few available in ambulatory basis.(17)

This study investigated the possible relationship between pre-existing conditions/comorbidities and onset symptoms with the risk of suffering poor outcomes in a population-based cohort of community-dwelling middle-aged and older adults with symptomatic COVID-19 (including hospitalised and ambulatory cases) in the region of Tarragona (old called Tarraco, Spain). In addition, we explored and developed simple COVID-19 screening rules to predict critical outcomes (ICU admission or death) useful in outpatient settings.

## METHODS

### Design, setting and study population

This is part of a large population-based retrospective-prospective cohort study (initiated in April 2020) involving 79,083 individuals in the region of Tarragona (Southern Catalonia, Spain). The design, setting and study population were extensively described in a prior article that evaluated the epidemiology of COVID-19 across the first wave of epidemic period in the study area.(18) This report focuses on community-dwelling individuals 50 years or older, who had a laboratory-confirmed COVID-19 (hospitalised or outpatient) in the study setting between March 1st to June 30th, 2020. The study was approved by the Ethical Committee of the Institution (Ethics Committee IDIAP Jordi Gol, Barcelona, file 20/065-PCV) and was conducted in accordance with the general principles for observational studies.(19) The study was determined to be exempt for informed consent under the public health surveillance exception.

### Data sources

The pre-existing CAPAMIS Research Database, an institutional clinical research database previously used for other epidemiological cohort studies in the area,(20) was updated for use as the main data source in this COVID-19 epidemiological investigation. Briefly, this research database compiles data from the institutional primary care clinical records electronic system (e-CAP), including administrative data and clinical information coded according to the International Classification of Diseases 10th Revision (ICD-10). It was used to identify sociodemographic characteristics and pre-existing conditions/comorbidities among cohort members and to establish their baseline characteristics at study start (01/03/2020).

When COVID-19 epidemic started, two electronic alerts including COVID-19’s laboratory registries plus ICD-10 codes for COVID-19 suspicion (B34.2, B97.29) were added to the e-CAP system and, later, both data sources were linked to construct the baseline research database used for the cohort study. In addition, in March 2020 a checklist of COVID-19 symptoms was quickly added to the e-CAP system to help family physicians in daily phone follow-up of COVID-19 patients. These electronic records, together complementary data registered during emergency visits and/or hospital-stay, were used to collect study data from patients. A trained research team of family physicians performed a retrospective review of the electronic health records to move the data to a standardised form, which was subsequently linked with the pre-existent main research database (which contained baseline characteristics of the 79,083 cohort members).

### Outcome definitions

A confirmed COVID-19 case was defined when a cohort member tested positive in SARS-COV-2 RT-PCR (reverse transcription-polymerase chain reaction) or in serological testing, according institutional guidelines.(21) A critical outcome was considered when the patient required ICU admission or died during clinical course. Death from COVID-19 was considered if the patient died from any cause within the first 30-day after the onset of the disease or in hospital-stay (any time).

### Covariates

Apart from demographic characteristics and pre-existing conditions/comorbidities (see Appendix), main covariates in this study were signs and symptoms of COVID-19 at presentation (within the first five days after the onset of illness). The check-listed symptoms (recruited according to data registered in the medical records, having been assessed by the attending physician in each case) were: cough, sore throat, rinorrhea (including runny nose and/or nasal congestion), fatigue, myalgias, headache, ageusia/disgeusia, anosmia, chest pain, dyspnea (shortness of breath), vomiting/nausea, diarrhea, confusion (including lethargy and/or delirium), general discomfort and fever (including low grade). A sign/symptom was considered absent if it was not recorded.

### Statistical analysis

Descriptive statistics of sociodemographic and clinical variables included frequencies, percentages, means and standard deviations (SD). Comparison of differences between groups was performed using Chi-squared or Fisher test for categorical variables and Student’s t test for continuous variables.

Logistic regression analyses were performed to calculate the Odds Ratios (crude, age/sex-adjusted and multivariable-adjusted ORs) that assessed the relationship between demographic characteristics, pre-existing comorbidities and signs / symptoms of COVID19 at presentation with the risk of suffering a critical outcome (ICU/death) during the course of the disease.

The criteria used to construct potential prognostic rules predicting critical outcome are described below. Developed rules included those covariables that showed significant or nearly significant association (p<0.10) with the dependent variable (ICU/death) in age/sex- and multivariable-adjusted logistic regression models. Discrimination and calibration were evaluated by means of receiver operating characteristic (ROC) curves, and the Hosmer-Lemeshow test, respectively. Sensitivity, specificity, positive and negative predictive values were calculated for distinct cut off points in the developed rules. In all analyses, statistical significance was set at p<0.05 (two-tailed). The analyses were performed using IBM SPSS Statistics for Windows, version 24 (IBM Corp., Armonk, N.Y., USA).

## RESULTS

Of the 345 overall laboratory-confirmed COVID-19 cases observed in non-institutionalised cohort members across the study period, 63 were excluded from this analysis because non-available clinical data (n=2), to be asymptomatic cases (n=27) or to be considered nosocomial cases occurred inside social-health hospital stay (n=34). Thus, the remaining 282 community-dwelling/non-institutionalised symptomatic COVID-19 cases were finally included in this report.

Mean age of cases was 65.9 years (SD: 12.1) and 140 (49.6%) were men. One hundred and fifty-four cases were hospitalised (30 of them were admitted in the ICU) and 128 were managed as outpatient (38 of them were attended in the emergency room but were not hospitalised). A total of 45 patients died (two in the emergency room before hospitalisation, 32 in hospital floor and 11 in the ICU). Overall, 64 (22.7%) suffered a critical outcome (ICU-admission or death).

The most prevalent comorbidities were hypertension (42.6%), obesity (25.5%), cardiac disease (23.4%), diabetes (19.1%) and respiratory disease (17.4%).

The most common signs/symptoms at COVID-19 presentation (first 5 days after onset of symptoms or medical contact) were fever (70.9%) and cough (67.7%) followed by general discomfort (42.2%), dyspnea (39.7%), fatigue (30.1%), myalgias (27.3%), diarrhea (24.1%) and headache (23%).

Table 1 compares demographic and clinical characteristics of cases according if they presented or not a critical outcome. Patients who suffered a critical outcome were older (mean age 73.6 vs 63.7 years, p<0.001) and had more comorbidities (2.6 vs 1.5 comorbidities, p<0.001) than those who did not suffer a critical outcome.

**Table 1.** Univariate analyses comparing characteristics (demographics, pre-existing comorbidities and early symptoms at presentation) in 282 community-dwelling COVID-19 patients over 50 years according to occurrence or not of a critical outcome (ICU-admission or death) across clinical course.

| Outcome<br>Variable | Non-ICU/death<br>(N=218)<br>n (%) | ICU/death<br>(N=64)<br>n (%) | p-value | Total<br>(N=282)<br>n (%) |
| --- | --- | --- | --- | --- |
| DEMOGRAPHICS |  |  |  |  |
| Age: 50-64 yrs | 142 (65.1) | 13 (20.3) | <0.001 | 155 (54.9) |
| 65-79 yrs | 56 (25.7) | 31 (48.4) |  | 87 (30.9) |
| ≥80 yrs | 20 (9.2) | 20 (31.3) |  | 40 (14.2) |
| Sex: Men | 101 (46.3) | 39 (60.9) | 0.040 | 140 (49.6) |
| Women | 117 (53.7) | 25 (39.1) |  | 142 (50.4) |
| COMORBIDITIES |  |  |  |  |
| Neurological | 4 (1.8) | 4 (6.3) | 0.061 | 8 (2.8) |
| Renal | 18 (8.3) | 11 (17.2) | 0.039 | 29 (10.3) |
| Cancer | 24 (11.0) | 10 (15.6) | 0.319 | 34 (12.1) |
| Rheumatic | 4 (1.8) | 1 (1.6) | 0.885 | 5 (1.8) |
| Respiratory | 28 (12.8) | 21 (32.8) | <0.001 | 49 (17.4) |
| Cardiac | 43 (19.7) | 23 (35.9) | 0.007 | 66 (23.4) |
| Atrial fibrillation | 16 (7.3) | 9 (14.1) | 0.096 | 25 (8.9) |
| Liver disease | 3 (1.4) | 3 (4.7) | 0.107 | 6 (2.1) |
| Hypertension | 83 (38.1) | 37 (57.8) | 0.005 | 120 (42.6) |
| Diabetes | 31 (14.2) | 23 (35.9) | <0.001 | 54 (19.1) |
| Smoking | 21 (9.6) | 4 (6.3) | 0.402 | 25 (8.9) |
| Alcoholism | 5 (2.3) | 2 (3.1) | 0.707 | 7 (2.5) |
| Obesity | 54 (24.8) | 18 (28.1) | 0.588 | 72 (25.5) |
| SYMPTOMS |  |  |  |  |
| Cough | 153 (70.2) | 38 (59.4) | 0.104 | 191 (67.7) |
| Sore throat | 31 (14.2) | 6 (9.4) | 0.313 | 37 (13.1) |
| Rinorrhea (RN) | 26 (11.9) | 2 (3.1) | 0.038 | 28 (9.9) |
| Myalgias | 73 (33.5) | 4 (6.3) | <0.001 | 77 (27.3) |
| Fatigue | 67 (30.7) | 18 (28.1) | 0.689 | 85 (30.1) |
| Headache | 53 (24.3) | 6 (9.4) | 0.010 | 59 (20.9) |
| Ageusia/disgeusia (AG) | 28 (12.8) | 3 (4.7) | 0.067 | 31 (11.0) |
| Anosmia (AN) | 34 (15.6) | 3 (4.7) | 0.023 | 37 (13.1) |
| Cluster RN/AG/AN | 61 (28.0) | 4 (6.3) | <0.001 | 65 (23.0) |
| Chest pain | 31 (14.2) | 6 (9.4) | 0.313 | 37 (13.1) |
| Dyspnea | 65 (29.8) | 47 (73.4) | <0.001 | 112 (39.7) |
| Vomiting | 12 (5.5) | 3 (4.7) | 0.798 | 15 (5.3) |
| Diarrhea | 53 (24.3) | 15 (23.4) | 0.886 | 68 (24.1) |
| Confusion | 5 (2.3) | 10 (15.6) | <0.001 | 15 (5.3) |
| General discomfort | 93 (42.7) | 26 (40.6) | 0.772 | 119 (42.2) |
| Fever | 154 (70.6) | 46 (71.9) | 0.849 | 200 (70.9) |

NOTE: Early symptoms were considered within the first five days of the onset of disease. p-values were calculated by using chi-squared or Fisher's test as appropriate.

In the crude analyses several pre-existing conditions/comorbidities (renal, respiratory, cardiac disease, diabetes and hypertension) and two symptoms (dyspnea and confusion) were significantly associated with a greater risk of admission to the ICU/death, whereas other symptoms (myalgias, headache and anosmia) were associated with a reduced risk. After age and sex-adjustment, apart from increasing age/years (OR: 1.07) and amle sex (OR: 1.64), only chronic respiratory disease (OR: 2.20) and diabetes (OR: 1.89), dyspnea (OR: 5.30) and confusion (OR: 4.95) remained significantly (or nearly significantly) associated with an increased risk, whereas myalgias (OR: 0.27) and the cluster symptom combining ageusia/anosmia/rinorrhea (or: 0.36) were associated with a significant reduced risk (table 2).

**Table 2.** Logistic regression analyses estimating crude and age/sex-adjusted Odds Ratios (Ors) to evaluate risks of suffering a critical outcome (ICU/death) by age, sex, pre-existing comorbidities and early symptoms in the 282 COVID-19 study subjects.

| Variable | UNADJUSTED<br>OR (95% CI) p | ADJUSTED<br>OR (95% CI) p |
| --- | --- | --- |
| **DEMOGRAPHICS** |  |  |
| Age, continuous yrs | 1.07 (1.05-1.10) <0.001 | 1.07 (1.05-1.10) <0.001 |
| Age group: 50-64 yrs | 1.00 (reference) <.001 | 1.00 (reference) <.001 |
| 65-79 yrs | 6.05 (2.95-12.39) <0.001 | 5.74 (2.79-11.82) <0.001 |
| ≥80 yrs | 10.92 (4.71-25.32) <0.001 | 10.59 (4.55-24.62) <0.001 |
| Sex male | 1.81 (1.02-3.19) 0.041 | 1.69 (0.92-3.11) 0.089 |
| **COMORBIDITIES** |  |  |
| Neurological | 3.57 (0.87-14.69) 0.078 | 1.91 (0.41-8.91) 0.411 |
| Renal | 2.31 (1.03-5.18) 0.043 | 1.05 (0.43-2.61) 0.912 |
| Cancer | 1.50 (0.68-3.32) 0.321 | 0.92 (0.38-2.19) 0.845 |
| Rheumatic | 0.85 (0.09-7.74) 0.885 | 0.78 (0.08-7.75) 0.830 |
| Respiratory | 3.31 (1.72-6.38) <0.001 | 2.20 (1.09-4.44) 0.028 |
| Cardiac | 2.28 (1.24-4.20) 0.008 | 1.11 (0.56-2.22) 0.762 |
| Atrial fibrillation | 2.07 (0.87-4.93) 0.102 | 0.83 (0.32-2.19) 0.712 |
| Liver disease | 3.53 (0.69-17.91) 0.129 | 2.43 (0.43-13.62) 0.312 |
| Diabetes | 3.38 (1.79-6.40) <0.001 | 1.89 (0.94-3.78) 0.073 |
| Hypertension | 2.23 (1.27-3.93) 0.006 | 1.06 (0.55-2.06) 0.855 |
| Smoking | 0.63 (0.21-1.89) 0.406 | 0.70 (0.21-2.33) 0.559 |
| Alcoholism | 1.37 (0.26-7.26) 0.708 | 1.28 (0.22-7.60) 0.784 |
| Obesity | 1.19 (0.64-2.22) 0.589 | 1.02 (0.52-2.00) 0.951 |
| **SYMPTOMS** |  |  |
| Cough | 0.62 (0.35-1.11) 0.106 | 0.68 (0.37-1.28) 0.234 |
| Sore throat | 0.62 (0.25-1.57) 0.316 | 0.79 (0.30-2.08) 0.634 |
| Rhinorrhea (RN) | 0.24 (0.06-1.03) 0.055 | 0.44 (0.10-1.97) 0.280 |
| Myalgias | 0.13 (0.05-0.38) <0.001 | 0.27 (0.09-0.81) 0.020 |
| Fatigue | 0.88 (0.48-1.63) 0.689 | 1.03 (0.53-2.00) 0.931 |
| Headache | 0.32 (0.13-0.79) 0.013 | 0.61 (0.24-1.60) 0.316 |
| Ageusia/disgeusia (AG) | 0.33 (0.10-1.14) 0.079 | 0.83 (0.22-3.08) 0.781 |
| Anosmia (AN) | 0.27 (0.08-0.90) 0.033 | 0.74 (0.20-2.72) 0.649 |
| Cluster AN/AG/RN | 0.17 (0.06-0.49) 0.001 | 0.36 (0.12-1.09) 0.070 |
| Chest pain | 0.62 (0.25-1.57) 0.316 | 0.69 (0.26-1.85) 0.464 |
| Dyspnea | 6.51 (3.48-12.17) <0.001 | 5.30 (2.75-10.24) <0.001 |
| Vomiting | 0.84 (0.23-3.09) 0.798 | 0.90 (0.23-3.59) 0.886 |
| Diarrhea | 0.95 (0.50-1.84) 0.886 | 1.14 (0.56-2.31) 0.713 |
| Confusion | 7.89 (2.59-24.04) <0.001 | 4.95 (1.49-16.43) 0.009 |
| General discomfort | 0.92 (0.52-1.62) 0.772 | 0.93 (0.51-1.70) 0.804 |
| Fever | 1.06 (0.57-1.97) 0.849 | 1.20 (0.62-2.36) 0.587 |

NOTE: Odds Ratios were calculated for those who had the condition as compared with those who had not it.

Finally, after multivariable-adjustment only age/years (OR: 1.04; 95% CI: 1.01-1.07; p=0.004), confusion (OR: 5.33; 95% CI: 1.54-18.48; p=0.008), dyspnea (OR: 5.41; 95% CI: 2.74-10.69; p<0.001) and myalgias (OR: 0.30; 95% CI: 0.10-0.93; p=0.038) remained significantly associated with increased or reduced risk of suffering a critical outcome (Table 3).

**Table 3.** Multivariable logistic regression model assessing risk of suffering a critical outcome (ICU-admission or death) among the 282 COVID-19 study subjects.

|  | **Odds Ratio** | **(95% CI)** | **p value** |
| --- | --- | --- | --- |
| **Age (continuous yrs)** | 1.04 | (1.01-1.07) | 0.004 |
| **Dyspnea** | 5.41 | (2.74-10.69) | <0.001 |
| **Confusion** | 5.33 | (1.54-18.48) | 0.008 |
| **Myalgias** | 0.30 | (0.10-0.93) | 0.038 |

NOTE: Odds Ratios were calculated for those who had the condition as compared with those who had not it. The Hosmer and Lemeshow test (Chi-square=5,683, with 8 df) was not significant (p= 0,683), indicating a good calibration of the model.

We used this multivariable-adjusted model to develop a simple predictive prognostic rule, named CD65-M (acronym for Confusion, Dyspnea, and Age >65 years, which contributed with a positive point each; and Myalgias which contributed with a negative point in the score), that showed an acceptable correlation with the risk of suffering a critical outcome (area under ROC curve: 0.828; 95% CI: 0.774-0.882).

Additionally, by using the logistic regression model adjusted by age and sex (table 2) we explored and developed a larger predictive rule including the 8 significant variables in that model. According to magnitude changes in the ORs, this rule called CD65RD-WMA (acronym of the initials of the names of the 8 included variables) weights 2 points for each one of Confusion and Dyspnea, 2 points for age greater than 80 years and one point for age between 65-79 years, one point for Respiratory disease and other for Diabetes. Furthermore, the score weights a negative point for each one of the following: Women, Myalgias and combined Ageusia/anosmia/rinorrhea. This longest CD65RD-WMA score (which may vary between −3 and 8 points) also showed a good correlation with the probability of suffering a critical outcome during clinical course, but did not substantially increased discriminatory power (area under ROC curve: 0,836; 95% CI: 0.784-0.889)

Figure 1 compares ROC curves for both CD65 and CD65RD-WMA rules.

**Figure 1.**
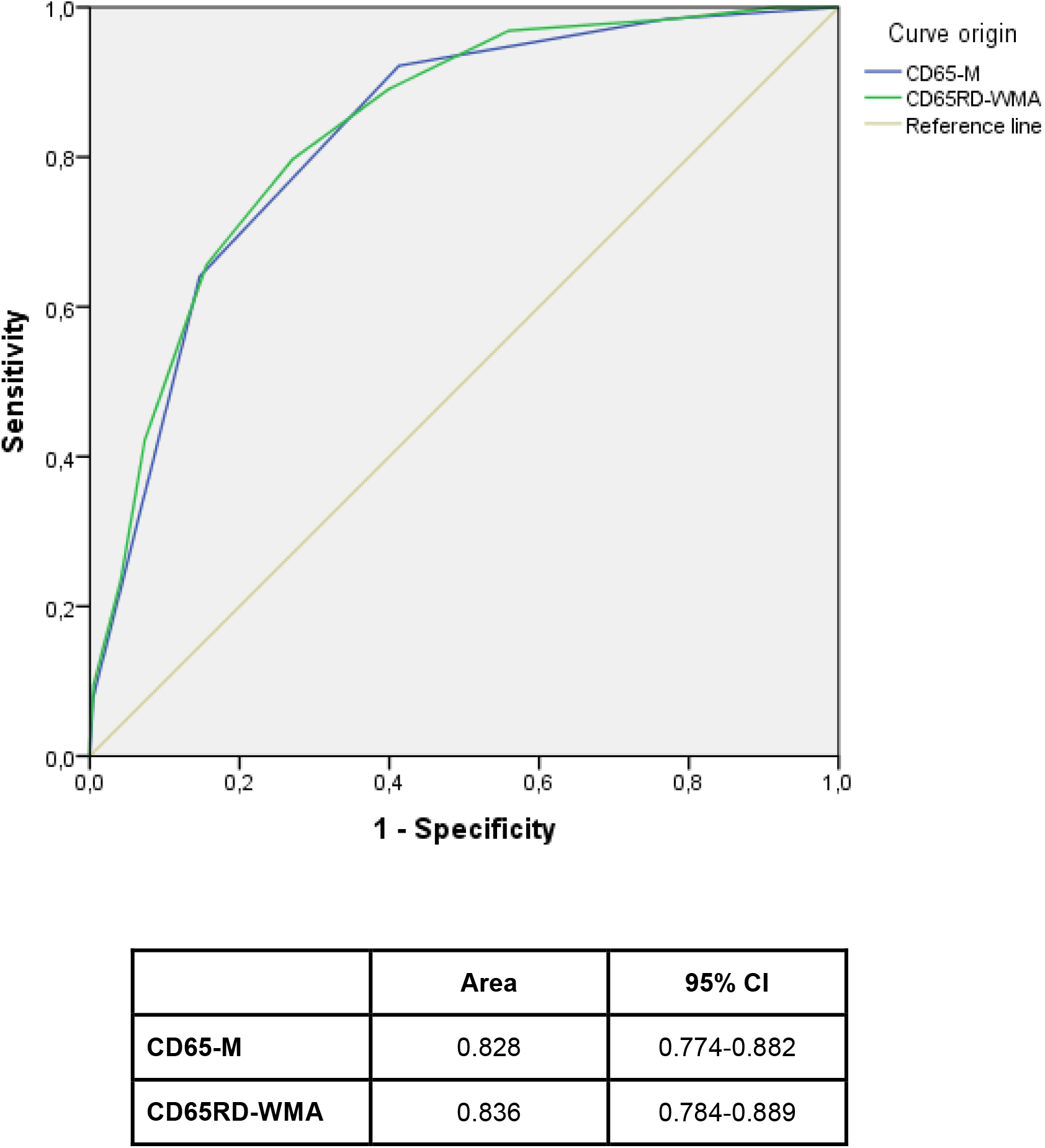
Comparison of ROC curves for both simple CD65-M and longer CD65RD-WMA rules.

Table 4 shows the absolute numbers and percentages of observed cases and critical outcomes in each scoring for both simple and longer rules.

**Table 4.**
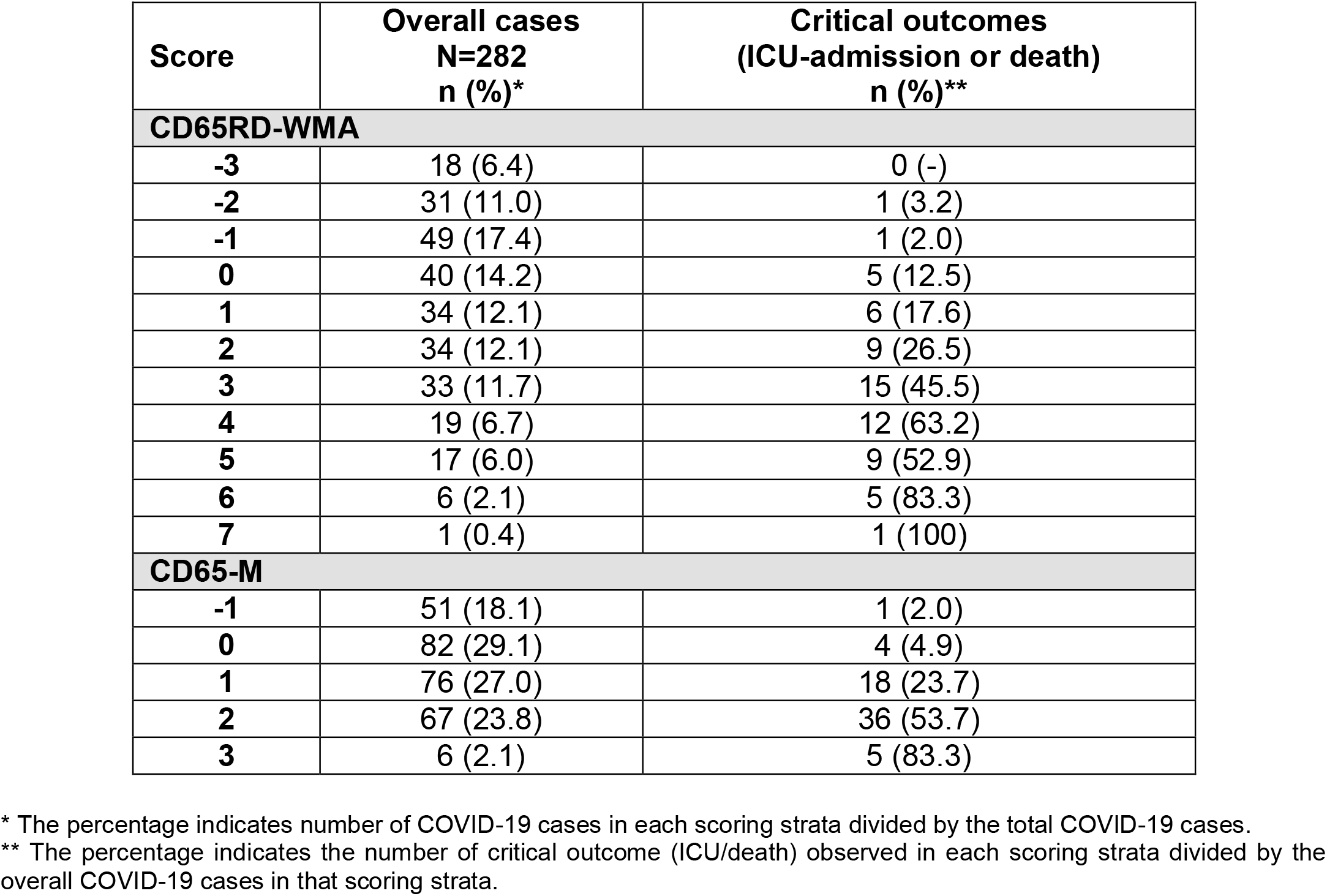
Distribution of overall COVID-19 cases and observed critical outcomes (ICU-admission or death) according to distinct scoring strata for both the simple CD65-M and the longer CD65RD-WMA prognostic rules.

Sensitivity, specificity, positive and negative predictive values for different cut off points in both rules are shown in Table 5.

**Table 5.**
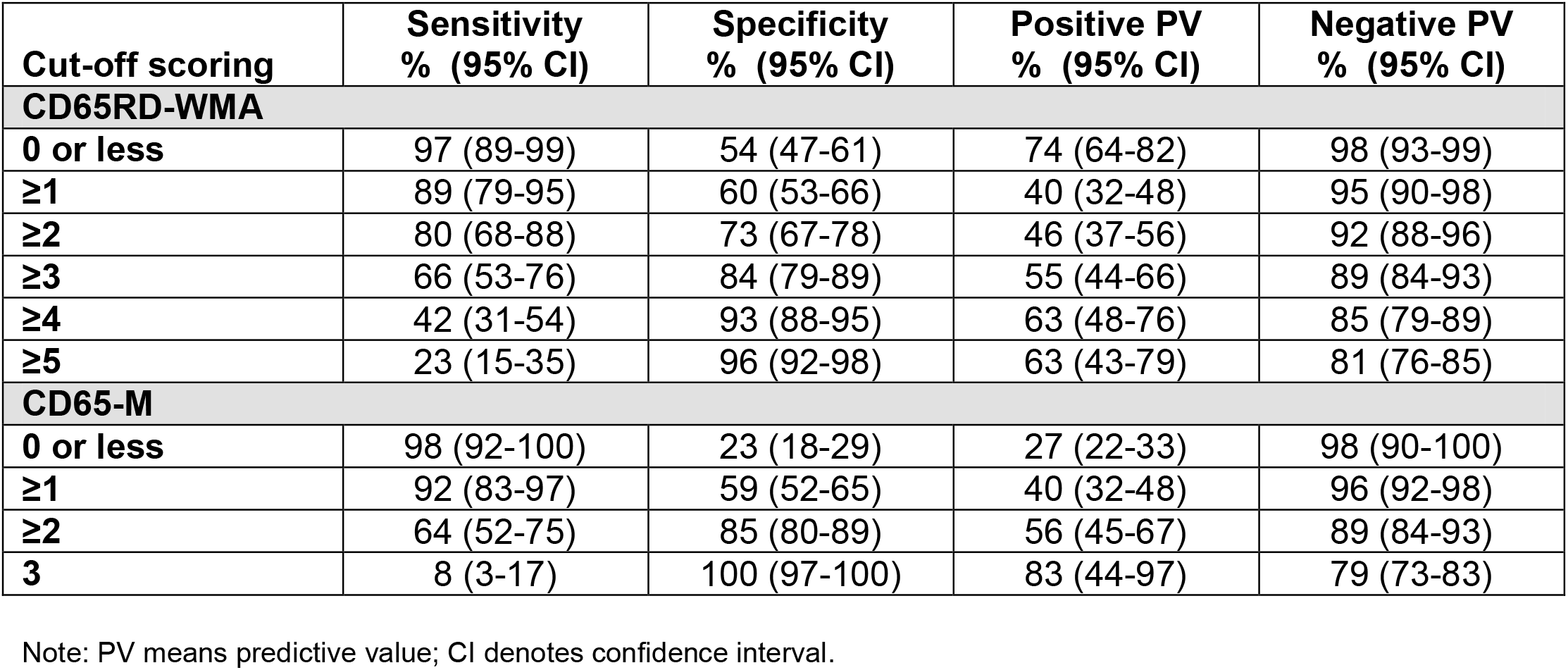
Sensitivity, specificity, positive and negative predictive values observed in the study population according to different cut off points for the CD65RD-WMA and CD65-M scores.

| **Cut-off scoring** | **Sensitivity**<br>**% (95% CI)** | **Specificity**<br>**% (95% CI)** | **Positive PV**<br>**% (95% CI)** | **Negative PV**<br>**% (95% CI)** |
| --- | --- | --- | --- | --- |
| **CD65RD-WMA** |  |  |  |  |
| **0 or less** | 97 (89-99) | 54 (47-61) | 74 (64-82) | 98 (93-99) |
| **≥1** | 89 (79-95) | 60 (53-66) | 40 (32-48) | 95 (90-98) |
| **≥2** | 80 (68-88) | 73 (67-78) | 46 (37-56) | 92 (88-96) |
| **≥3** | 66 (53-76) | 84 (79-89) | 55 (44-66) | 89 (84-93) |
| **≥4** | 42 (31-54) | 93 (88-95) | 63 (48-76) | 85 (79-89) |
| **≥5** | 23 (15-35) | 96 (92-98) | 63 (43-79) | 81 (76-85) |
| **CD65-M** |  |  |  |  |
| **0 or less** | 98 (92-100) | 23 (18-29) | 27 (22-33) | 98 (90-100) |
| **≥1** | 92 (83-97) | 59 (52-65) | 40 (32-48) | 96 (92-98) |
| **≥2** | 64 (52-75) | 85 (80-89) | 56 (45-67) | 89 (84-93) |
| **3** | 8 (3-17) | 100 (97-100) | 83 (44-97) | 79 (73-83) |

Note: PV means predictive value; CI denotes confidence interval.

## DISCUSSION

In the current context of global pandemic and public health emergency by COVID-19, the knowledge of protective and/or risk factors to suffer infection or develop poor outcomes is essential for adequate management of patients in earlier stages of the disease.(17) In the present community-based cohort study, apart from increasing age that has repeatedly documented as major risk condition,(4-8) we have also found that some early symptoms (within the first five days after the onset of disease) are associated with better or poor prognosis (i.e, need of ICU admission or death). Indeed, presence of dyspnea and confusion clearly increased the risk of ICU-admission or death, whereas myalgia was associated with a reduced risk.

On the first, it is not surprising since both variables, or related-variables, were already associated with an increased risk of death in patients with community-acquired pneumonia, having been already included confusion and taquipnea as predictor variables within simple prognostic rules (such as CURB-65 or CRB-65 rules) assessing severity in patients with pneumonia.(22-24)

On the second, surprisingly, myalgias emerged independently associated with a reduced risk for critical outcomes after multivariable adjustments. This may be considered as an unexpected result, but we note that it is not unique and has already been reported by other studies. Concretely, in a single-centre French cohort study that included 150 hospitalised COVID-19 consecutive patients between March-April 2020, the percentage of cases who had myalgias was significantly greater among those patients with favourable clinical course than in those who required ICU-admission or died.(16)

Thus, we constructed a simple prognostic rule containing the above mentioned four variables (CD65-M score), which showed a good correlation with the probability of suffering a critical outcome across clinical course, being 2%, 4.9%, 23.7%, 53.7% And 83.3% for scoring −1, 0, 1, 2 and 3, respectively. According this CD65-M rule, the best cut-off point for predicting bad evolution would be scoring ≥1 (92% sensitivity with 59% specificity) or scoring ≥2 (64% sensitivity with 85% specificity).

If we consider pre-existing comorbidities, many studies have reported increased risk of poor outcomes among patients with diverse comorbidities (especially cardiac, respiratory or renal disease, diabetes, obesity and hypertension), but most studies assessed crude associations (without age- and/or multivariable-adjusted risk).(4-8,25-29)

In the present study we also observed these associations in the crude analyses, but after multivariable adjustment none pre-existing comorbidity was associated with an adjusted increased risk in our study population.

Nevertheless, since respiratory disease and diabetes were significantly associated with an increased risk in the age and sex-adjusted model, we explored and developed a second lengthy rule (CD65RD-WMA) including significant variables in that model. However, this second model did not substantially improve predictive performance of the first simpler CD65-M rule (containing exclusively the 4 significant predictor variables after multivariable-adjustment).

Main strengths in this study were its population-based design (including as hospitalised as well as ambulatory COVID-19 cases), the assessment of early COVID-19 symptoms as possible predictors (rare in the literature), and the large follow-up necessary to adequately recruit the occurrence of critical outcomes during the clinical course (i.e, deaths occurred in hospitalised COVID-19 patients several weeks after hospital or ICU admission). In addition, we performed multivariable-adjusted analysis to accurately estimate relationships between baseline covariables and risk of suffering a critical outcome, developing simple predictive prognostic rules (which may be especially useful in primary care settings). The observed prevalence for most symptoms fits with data reported in other epidemiological studies,(30) which supports the validity of collected data despite retrospective design.

As major limitations, our study was conducted in a single geographical area with specific epidemic conditions (relatively low incidence of COVID-19) in a limited time period (first wave of pandemic) and focused exclusively on community-dwelling people over 50 years.(18) We don’t know the possible influence that changes in study population and/or epidemic intensity could have on the statistics reported here. We also note that the relatively little sample size (282 cases with 64 outcomes in our study) building prediction models may increase the risk of overfitting the model, which implies that the performances of the models in new samples could be worse. We underline that a further validation in an external cohort is necessary before a routine use of the described prognostic rules may be applied. We note, however, that prediction models are needed to support medical decision managing COVID-19 patients.

There are several published or preprint reports that developed prognostic rules predicting critical outcomes (need of mechanical ventilation and/or death), but all of them were based on complementary explorations or laboratory findings which are not generally available in ambulatory or primary care settings.(17) Main contribution of this study lies in the fact that it reports simple clinical prognostic rules including easily accessible clinical data (such as demographics, pre-existing comorbidities and early symptomatology), which could be very helpful assessing COVID-19 patients outside the hospital.

We conclude that the clinical course of COVID-19 patients remains unpredictable in earlier stages of the disease, but simple clinical tools as the proposed CD65-M prognostic rule (pending external validation) may be helpful assessing these patients in primary care settings. Further evaluations adding other easily accessible complementary data (e.g,, pulse-oximetry to detect silent hypoxia) could improve predictive performance.

## Data Availability

These data have been obtained from the Catalonian Health Institute Information System for the Development of Research in Primary Care (SIDIAP). Interested authors might obtain SIDIAP data (previous ethics and scientific approval by the ethics and clinical research committee of the Primary Care Research Institute Jordi Gol (IDIAP Jordi Gol)) addressing purposes to the Institution.

## ACKNOWLEDGEMENTS

This study was supported by a grant from the Instituto de Salud Carlos III of the Spanish Health Ministry (file COV20/00852; call for the SARS-COV-2/COVID-19 disease, RDL 8/2020, March 17, 2020).

## CONTRIBUTORS

AVC designed the study; AVC and ESG assessed outcomes and wrote the manuscript; ESG, CDC, MFP and IHG obtained data; ESG and AVR did statistical analyses and edited the manuscript; OOG revised the final version; AVC coordinated the study.

## APPENDIX. Criteria used to identify pre-existing conditions/comorbidities in the study population

**APPENDIX. Criteria used to identify pre-existing conditions/comorbidities in the study population.**
| Comorbidities with ICD-10 codes [International Classification of Diseases, 10th Revision] |  |
| --- | --- |
| **Neurological disease:**<br>Dementia<br>Ictus | F01-F03<br>I63, I61 |
| **Chronic renal failure** | N18-N19 |
| **Cancer** (solid organ or haematological neoplasia) in past 5 years | C00-C97 |
| **Rheumatologic disease:**<br>Rheumatoid arthritis, enteropathic arthropathies and juvenile arthritis<br>Systemic lupus erythematosus | M05-M09<br>M32 |
| **Chronic pulmonary/respiratory disease:**<br>Chronic bronchitis/emphysema<br>Asthma<br>Other chronic pulmonary diseases | J41-J44<br>J45-J46<br>P27, E84, J47 |
| **Chronic heart disease:**<br>Congestive heart failure<br>Coronary artery disease<br>Other chronic heart diseases | I50<br>I20-I22, I25<br>I05-I08, I11, I35-I37, I42, I51.7 |
| **Atrial Fibrillation** | I48 |
| **Chronic liver disease:**<br>Chronic viral hepatitis<br>Cirrhosis<br>Alcoholic hepatitis | B18<br>K74<br>K70 |
| **Diabetes mellitus** | E10-E14 |
| **Hypertension** | I10, I11, I12 o I15 |
| **Hypercholesterolemia** | E78 |
| **Obesity** | E66 |
| **Smoking** | F17 |

